# Developing A Data Pipeline to Quantify Ventilator Waveforms

**DOI:** 10.1101/2025.10.28.25339000

**Authors:** Peter D Sottile, Lenny Larchick, J.N. Stroh, David Albers, Bradford Smith

## Abstract

**Objective:** To automatically collect ventilator waveforms and integrate them with curated electronic health record data from thousands of patients to provide the data necessary to analyze the complex interactions between lung injury, patient effort, ventilator dyssynchrony, and ventilator mechanics. Such datasets do not currently exist, hampering the understanding of ventilator trajectories.

**Design:** A prospective, observational study which utilizes a multidisciplinary team of data scientists, biomedical engineers, and clinicians to develop an automated pipeline collecting high-fidelity ventilator waveform data and integrating these data with electronic health record data, including vital signs, sedation and agitation scores, lab results, and medications (drug, dose, and route). Importantly, electronic health record data are collected over a patient’s entire hospital course, allowing for a complete description of patient trajectories.

**Settings:** The Medical Intensive Care Unit at the University of Colorado.

**Patients:** All mechanically ventilated adult patients.

**Interventions:** None.

**Measurements:** Automated collection of high-fidelity ventilator waveforms and electronic health record data.

**Results:** Between July 2023 and May 2025, we collected data from 1,116 patients, 968 (87%) of whom had >12 hours of mechanical ventilation. These patients generated 4,767 ventilator days (>13 ventilator years) of analyzable ventilator waveforms and had a median duration of ventilation of 2.6[1.25, 6.06] days. Over 146 million breaths were segmented from the waveforms, of which 49 million breaths were able to fit the linear single-compartment model accurately and had a median compliance of 35.7 [25.2, 45.3] mL/H_2_O. Electronic health record data was linked to the waveforms to provide 8,511 [3,835, 17,040] records per patient. These data constitute a comprehensive database for studying the effects of mechanical ventilation, patient effort, ventilator dyssynchrony, and key non-ventilator covariates, such as sedation, across a large and heterogeneous cohort of patients.

**Conclusion:** We created a fully automated data pipeline to continuously collect mechanical ventilation waveform data and integrate it with detailed EHR data to generate a unique, high-fidelity dataset that will be crucial for understanding the complex relationships among lung injury, patient effort, sedation, ventilator dyssynchrony, and ventilator mechanics.

**Key Points:** *Question:* To create an automated data pipeline to collect and integrate continuous ventilator waveform data with electronic health record data.

*Findings:* Between July 2023 and May 2025, we automatically collected data from 1,116 patients, 968 (87%) of whom received mechanical ventilation for more than 12 hours.

*Meaning:* This fully automated data collection pipeline will facilitate advances in understanding the complex relationships between lung injury, patient effort, sedation, ventilator dyssynchrony, and ventilator mechanics.

## Introduction

Invasive mechanical ventilation is a life-saving intervention. Continuous ventilator waveforms of airway pressure, flow, and volume are routinely displayed on modern ventilators, providing detailed diagnostic and prognostic information at the bedside related to lung physiology and injury, patient respiratory effort, ventilator dyssynchrony, and ventilator mechanics, which can rapidly change over time.(1) However, systematic and longitudinal analysis of these data have historically been limited to a few descriptive variables, such as tidal volume, peak inspiratory pressure, or respiratory rate. Such variables are recorded relatively infrequently in the electronic health record (EHR) and inadequately capture the feature-rich, high-fidelity waveform data observed at the bedside.

Importantly, mechanical ventilation can injure the lungs.(2) Ventilator-induced lung injury (VILI) is caused by increases in lung strain (approximated as volumetric lung deformation) and stress (approximated as the force per unit area or transpulmonary pressure).(3, 4). Low-tidal volume ventilation is one strategy to decrease lung strain and stress in patients, thereby improving outcomes in patients both at risk for and with acute respiratory distress syndrome (ARDS).(5–9) However, low-tidal volume ventilation does not always prevent VILI, especially in the presence of respiratory efforts.(10– 13) Indeed, ventilator dyssynchrony is defined as the inappropriate timing and delivery of a ventilator breath in response to respiratory effort and is associated with increased markers of lung strain and stress, as well as increased mortality.(14–21) Consequently, to understand the complex interactions between lung injury, patient effort, ventilator dyssynchrony, sedation, and ventilator mechanics, it is essential to analyze complete waveforms of each breath, as details related to lung mechanics, patient effort, and ventilator dyssynchrony are not well captured in traditional descriptive electronic health record (EHR) data.

Such detailed data have been unavailable for analysis due to several reasons. First, the average patient receiving mechanical ventilation receives over 25,000 breaths daily. Historically, nearly 40% of intensive care unit (ICU) admissions require mechanical ventilation, resulting in millions of mechanically ventilated breaths daily in a single hospital.(22) To recapitulate the details of airway pressure, flow, and volume waveforms, data must be captured at a sufficient resolution, typically 30-50 Hz for each waveform, over the entire course of mechanical ventilation (typically several days). Moreover, to fully understand a patient’s clinical course, data must be collected throughout their entire duration of mechanical ventilation. For instance, snapshots of data miss short but critical events (i.e., clusters of dyssynchrony) that define a patient’s clinical trajectory, while the development and resolution of VILI occurs over days.(23) The technology to capture, transmit, and store these data has only recently been introduced to the ICU, generating tremendous amounts of data that healthcare systems have traditionally been ill-equipped to collect.

Second, elucidating meaningful interpretations of these waveform data throughout mechanical ventilation requires the development of automated analysis pipelines. A single patient generates approximately 25,000 breaths in a day, which rapidly scales to millions when studying hundreds or thousands of patients. Tasks range from the seemingly simple to the complex, including 1) extracting waveform data from different ventilators, 2) securely transmitting that data to secure servers, 3) transforming waveform data into usable data streams, 4) segmenting continuous waveforms into individual breaths, 5) aligning timestamps from different sources (ventilator and EHR), and 6) developing methods to analyze individual breaths, including the creation and validation of automated machine learning algorithms to identify dyssynchrony and quantify critical features observed in ventilator waveforms.(15, 24–33) Only in the last decade has the computational capacity to handle such tasks become readily available.

Third, waveform data only partially describes the respiratory system and lacks the context of the patient’s condition, treatments, and outcomes. Ventilation strategies often require a delicate balance between optimizing respiratory targets, minimizing sedation, and ensuring adequate hemodynamics. In other words, the ventilator is not utilized in isolation from the rest of the patient. Consequently, accurately interpreting ventilator waveform data requires quantifying the broader context of the patient’s condition. To accomplish this, ventilator waveform data must be integrated into the larger EHR, including vital signs, sedation and agitation scores, lab results, positioning, fluid balance, and medication strategies (e.g., sedation).

Finally, developing such pipelines requires a multidisciplinary team of data scientists, biomedical engineers, health system information technology (IT) experts, and clinicians to build, test, and validate the end-to-end data pipeline. Integrating waveform pipelines with EHR data requires motivated and persistent individuals with expertise in wrangling incomplete and “messy” EHR data. Building such teams has only recently become a priority for healthcare systems.

### Objective

The inability to automatically capture ventilator waveform data and integrate it with curated EHR results *throughout a patient’s entire hospital encounter* impairs our understanding of the complex interactions between lung injury, patient effort, ventilator dyssynchrony, sedation strategies, and ventilator mechanics. Prior data are limited to short durations of mechanical ventilation or lack integration with the EHR to obtain key non-ventilator covariates. Large, heterogeneous databases with such data do not exist for analysis. Moreover, building such a pipeline is the first step in leveraging these data streams for real-time clinical utilization by developing, testing, and validating predictive models to inform clinical decision-making.

Consequently, we sought to develop a secure computational pipeline to automatically collect high-fidelity ventilator waveform data on all mechanically ventilated patients in the Medical Intensive Care Unit (MICU) at the University of Colorado, a quaternary-care university hospital. We integrated these data with EHR data, including vital signs, sedation and agitation scores, lab results, and medications (drug, dose, and route) from the entire hospitalization. This pipeline conforms to modern data exchange standards (Health Level Seven International (HL7)) and regulatory requirements (Health Insurance Portability and Accountability Act (HIPAA)), satisfying both US regulatory requirements and international interoperability expectations.(34–36) Previously, such a dataset did not exist to aid ventilator research. This study presents a computational pipeline and describes the dataset collected to facilitate future studies aimed at reducing ventilation-related morbidity and mortality, providing insight into the effects of ventilator dyssynchrony and key non-ventilatory covariates, such as sedation, in a large and heterogeneous patient cohort.

## Materials and Methods

### Patients Included

The data pipeline was developed at the UCHealth University of Colorado Hospital to capture data from all patients admitted to the Medical Intensive Care Unit (MICU) who receive mechanical ventilation. The UCHealth University of Colorado Hospital is a multi-specialty, quaternary hospital with over 800 beds and 2.6 million annual patient visits. The MICU provides sub-specialty care for adult patients, ages greater than 14 years old, with a variety of critical illnesses, including septic shock, ARDS, and general respiratory failure (i.e., pneumonia, asthma, interstitial lung disease (ILD), chronic obstructive pulmonary disease (COPD)), gastrointestinal hemorrhage, and a variety of oncological disorders. The MICU does not routinely care for postoperative patients or patients with primary cardiac pathology.

Patients are ventilated in the MICU with either a Hamilton G5 or a C6 ventilator (Hamilton Medical AG, Switzerland). This data pipeline was approved by the University of Colorado Combined Institutional Review Board (COMIRB, #20-2160) with waived consent, allowing the collection of clinical waveform and EHR data from all mechanically ventilated patients.

### Development of A Data Model

Ventilator data, including continuous airway pressure, flow, and volume measured at 50 Hz, are included to recapitulate ventilator waveforms. Ventilator settings are captured once per minute, including set ventilator mode, tidal volume, inspiratory pressure, positive end-expiratory pressure (PEEP), rate, inspiratory to expiratory time ratio (I:E ratio), rise time, flow pattern, and other mode-specific settings (i.e., high time (t-high) if the patient was in an airway pressure release ventilation (APRV) mode of ventilation). EHR data included patient age, sex, race, ethnicity, vital signs (i.e, temperature, blood pressure, heart rate, observed respiratory rate, pulse oximetry), physiologic data (i.e, Richmond Agitation and Sedation Score (RASS), hourly urine output, patient positioning, and nursing and respiratory therapist assessments), all laboratory test results, medication administration times, route, and dosages (i.e., vasopressors, sedatives, anxiolytics, and neuromuscular blockage agents), in-hospital transfers between units, and outcomes (i.e., discharge location and mortality). These data were deemed necessary by the clinical team to understand the effects of mechanical ventilation, especially ventilator dyssynchrony, and key non-ventilator covariates, such as sedation, on a large and heterogeneous patient cohort.

### Waveform Data Collection

We established a team within UCHealth comprising data scientists, biomedical engineers, health system information technology (IT) experts, and clinicians to collect high-fidelity waveform data as outlined in Figure 1. Each bed in the MICU uses the Capsule Neuron 3 (Philips, Cambridge, MA) to automate data collection from accessory devices, including IV pumps, dialysis machines, and mechanical ventilators. The Capsule management software is part of the on-site hospital computing infrastructure, running on a local virtual machine (VM) (VM1 in Figure 1). This software allows for the management of the Neurons (including updating software and changing settings), facilitates the translation of proprietary ventilator data streams to HL7, and has ‘output connectors’ for transferring the data to the desired data sink. The Capsule management software sends clinical production data to the local Epic instance (Epic, Verona, WI, US) and the research waveform data through our research pipeline. The details of our research data pipeline, including data transfer, transformation, breath segmentation, and pipeline monitoring, are described in Appendix I: Waveform Data Processing.

**Figure 1:**
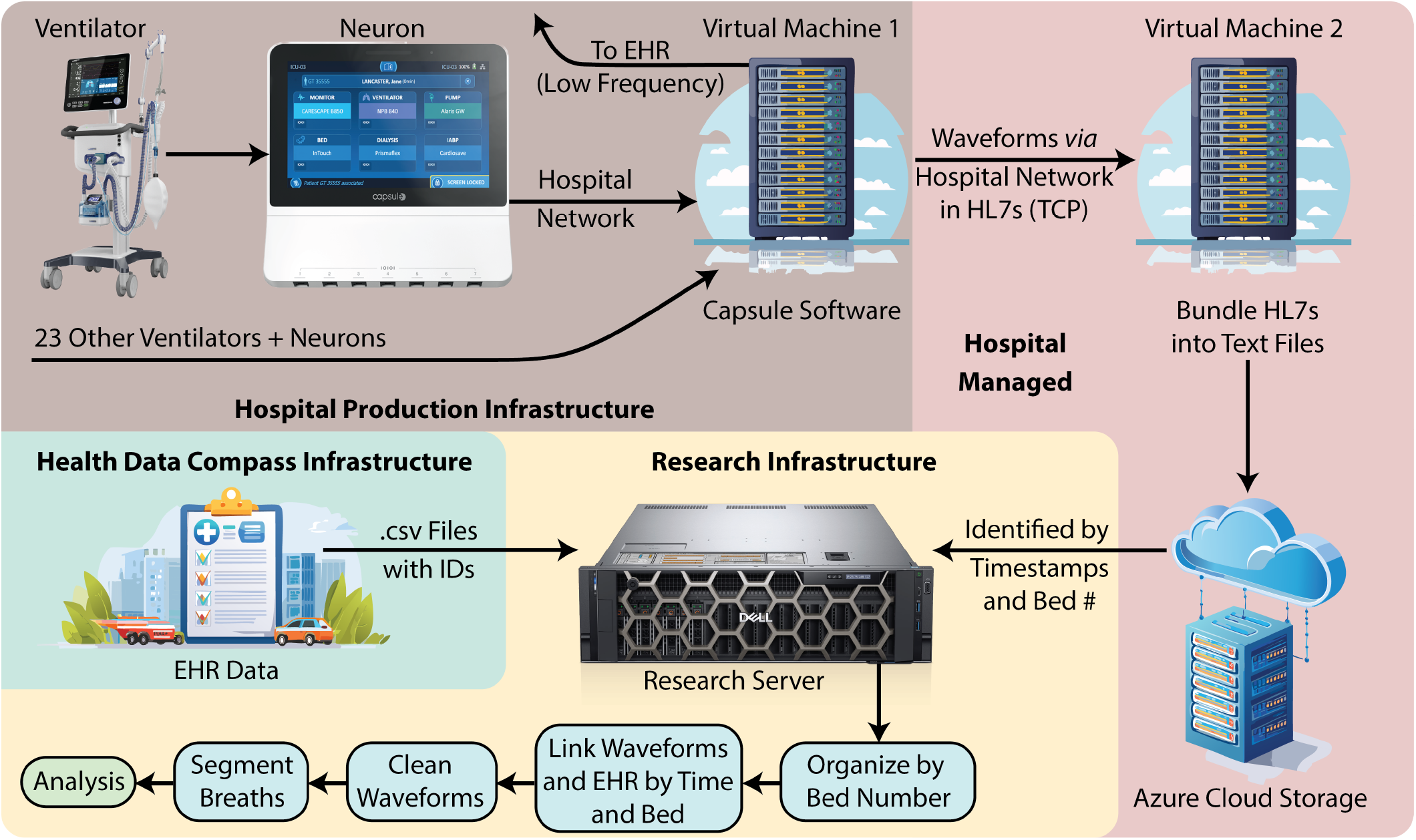
Data pipeline flowchart: Ventilator waveform data travels through the Capsule Neuron in-room devices to a management server (Virtual Machine 1). This pathway shares infrastructure with the production EHR pipeline. The data are then sent to Virtual Machine 2, where they are aggregated and uploaded to an Azure Cloud Storage instance. This infrastructure is hospital-managed but not part of the production EHR pipeline. Our research server downloads those waveforms once daily for the Azure Cloud. EHR data is provided by Health Data Compass on request and linked to the waveforms using bed occupancy.

### Ehr Integration

We leverage Health Data Compass (HDC) (healthdatacompass.org), a multi-institutional health data warehouse at UCHealth, to link time-stamped patient-specific EHR data to the ventilator waveform data. Patient data is integrated into the HDC enterprise data warehouse through processes developed by software engineers to extract data from various source systems, transform that data into a standard schema (OMOP CDM V5), and load the data into the data warehouse. Relevant administrative and clinical data are then extracted into data marts. The details of our research EHR data pipeline are described in Appendix II: EHR Integration.

### Regulatory Compliance

This data pipeline was primarily designed to maintain HIPAA and HL7 compliance. Importantly, HDC data analysts provide institutional honest-broker services for investigators requiring access to limited data sets, de-identified data sets, or data marts, which are critical to this project. The Colorado Clinical and Translational Sciences Institute (CCTSI) ensures the HIPAA compliance of the research computational server.

### Waveform Analysis

Basic individual-breath descriptive variables, such as peak pressure, peak flow, delivered tidal volume, respiratory rate, and observed PEEP, are readily calculated from the raw waveform data. The continuous waveform data also allows for the calculation of more advanced metrics. For example, the linear single compartment model (SCM) model was efficiently fit to each breath using matrix inversion, yielding accurate estimates of resistance and compliance independent of specific ventilator maneuvers performed at the bedside to measure these values.(37) Moreover, continuous mechanical power was directly calculated as the time-based integral of the pressure flow product for each breath.

Patient characteristics or other EHR derived variables can then stratify these data. To demonstrate the power of these data, we utilized International Classification of Diseases, 10th revision (ICD-10) codes to identify patients with ARDS or ARDS risk factors, as previously described. We compared them with patients with other causes of respiratory failure.(38–40) Values were compared with logistic regressions for binomial tests, Wilcoxon rank sum for non-normally distributed variables, t-tests for normally distributed variables, and mixed-effects models to account for repeated measurements within a patient for breath characteristics. Importantly, these data lend themselves to much more nuanced, time-based analysis.

## Results

This rich, unique data pipeline yields a substantial amount of harmonized data detailing the entire patient encounter. Between July 2023 and May 2025, we collected data from 1116 patients, 968 (87%) of whom had received mechanical ventilation for more than 12 hours. Of these patients, 704 (72%) have ARDS or ARDS risk factors as defined by ICD-10 codes, and 421 were female (43%); demographics are detailed in Table 1. Patients had an average age of 55.2 ± 17.4 and an in-hospital mortality of 33.8%.

**Table 1:**
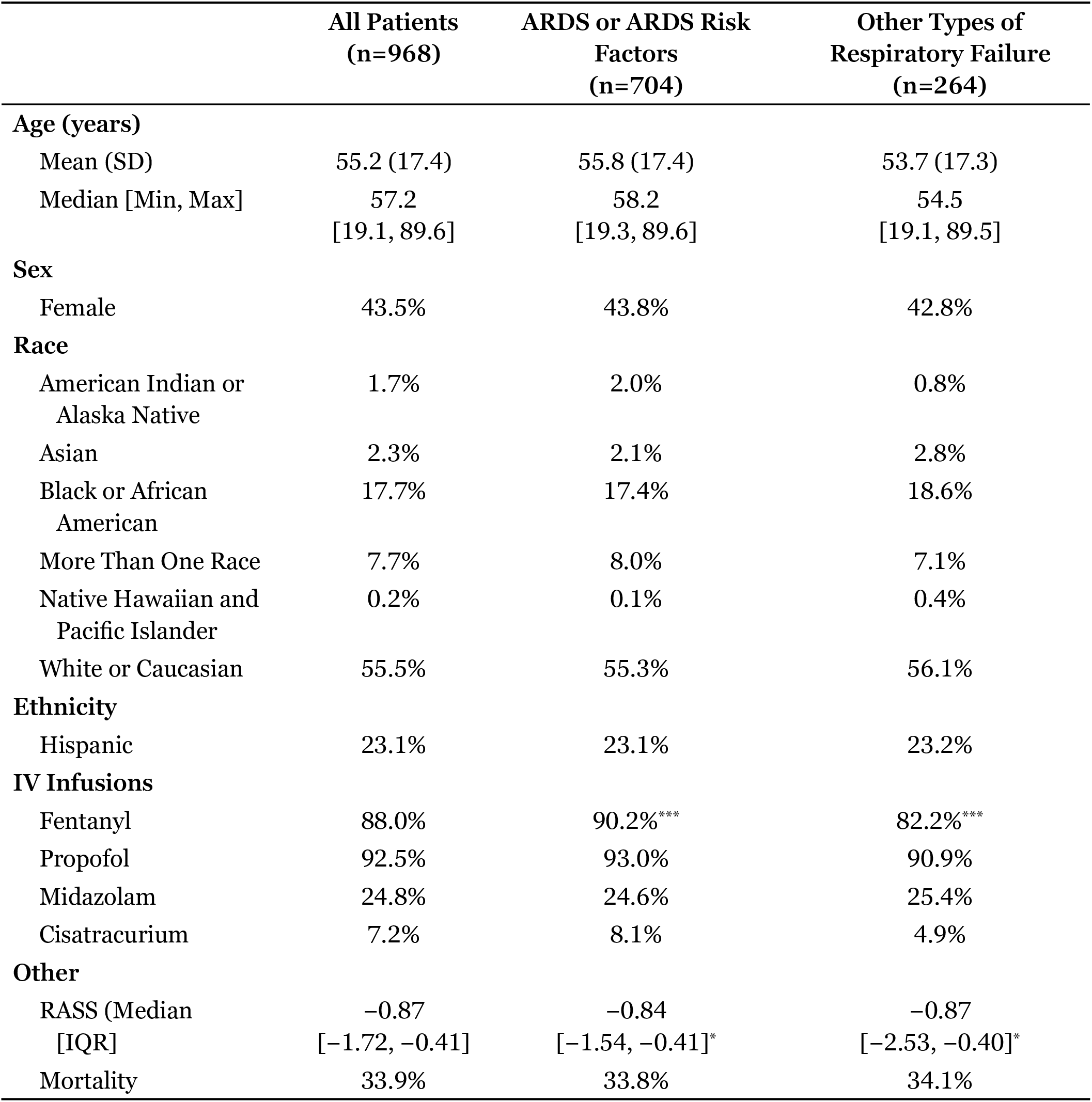
Descriptive characteristics (demographics, IV infusions for analgesia, sedation, and neuromuscular blockade, RASS, and mortality) of patients captured by the data pipeline between July 2023 and May 2025 and stratified by patients with ARDS or ARDS risk factors compared to other patients. ^*^ p < 0.05, ^***^ p < 0.005

|  | All Patients<br>(n=968) | ARDS or ARDS Risk<br>Factors<br>(n=704) | Other Types of<br>Respiratory Failure<br>(n=264) |
| --- | --- | --- | --- |
| **Age (years)** |  |  |  |
| Mean (SD) | 55.2 (17.4) | 55.8 (17.4) | 53.7 (17.3) |
| Median [Min, Max] | 57.2<br>[19.1, 89.6] | 58.2<br>[19.3, 89.6] | 54.5<br>[19.1, 89.5] |
| **Sex** |  |  |  |
| Female | 43.5% | 43.8% | 42.8% |
| **Race** |  |  |  |
| American Indian or<br>Alaska Native | 1.7% | 2.0% | 0.8% |
| Asian | 2.3% | 2.1% | 2.8% |
| Black or African<br>American | 17.7% | 17.4% | 18.6% |
| More Than One Race | 7.7% | 8.0% | 7.1% |
| Native Hawaiian and<br>Pacific Islander | 0.2% | 0.1% | 0.4% |
| White or Caucasian | 55.5% | 55.3% | 56.1% |
| **Ethnicity** |  |  |  |
| Hispanic | 23.1% | 23.1% | 23.2% |
| **IV Infusions** |  |  |  |
| Fentanyl | 88.0% | 90.2%*** | 82.2%*** |
| Propofol | 92.5% | 93.0% | 90.9% |
| Midazolam | 24.8% | 24.6% | 25.4% |
| Cisatracurium | 7.2% | 8.1% | 4.9% |
| **Other** |  |  |  |
| RASS (Median<br>[IQR]) | -0.87<br>[-1.72, -0.41] | -0.84<br>[-1.54, -0.41]* | -0.87<br>[-2.53, -0.40]* |
| Mortality | 33.9% | 33.8% | 34.1% |

These patients generated 4,767 ventilator days (>13 ventilator years) of analyzable ventilator waveforms and had a median duration of ventilation of 2.6 [IQR 1.25, 6.06] days. Waveform data generated 146 million individual breaths. Our rule-based segmentation algorithm achieved an accuracy of over 97.1% in matching the start and stop points of breaths within 0.1 seconds, compared with the approach used by the Hamilton DataLogger software, which identifies when the ventilator’s inspiratory and expiratory valves open and close. Equally important, our pipeline monitoring tools improved and ensured the collection of complete data, as demonstrated in Figure 2.

**Figure 2:**
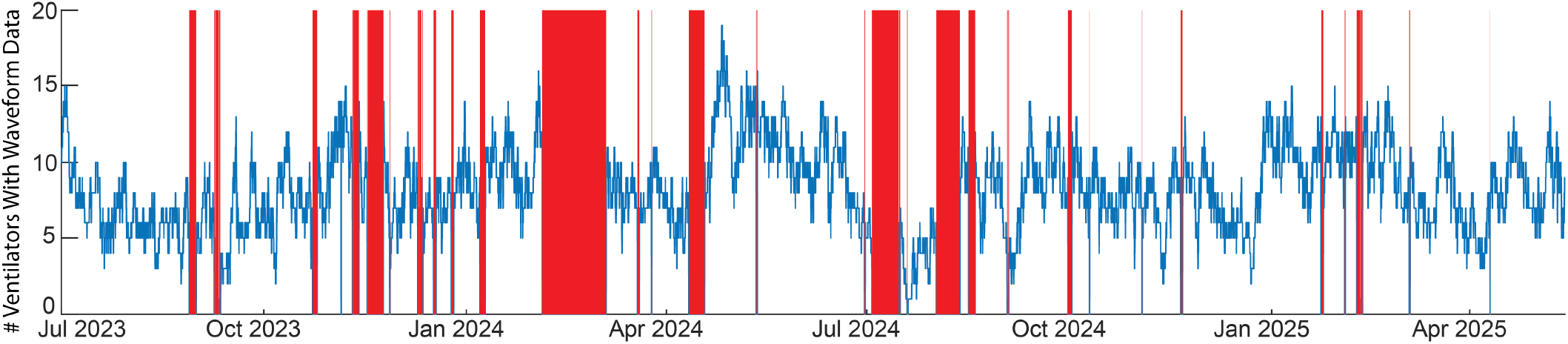
Ventilator Utilization and Data Pipeline Uptime: Time series of the number of simultaneous MICU ventilator waveforms collected over time (blue). Red bands show pipeline outages.

The linkage of EHR to waveform data was effective, and 98.3% of 2024 waveforms were assigned to a patient with waveform data, as shown in Figure 3. Temporal alignment of EHR and waveform data was checked by comparing the waveform-estimated and EHR-reported PEEP. The median difference was −0.06 [−0.25, 0.03] cmH_2_O, and inspection of breaths with substantial errors shows many instances of respiratory efforts pulling the waveforms below the set (EHR-reported) PEEP.

**Figure 3:**
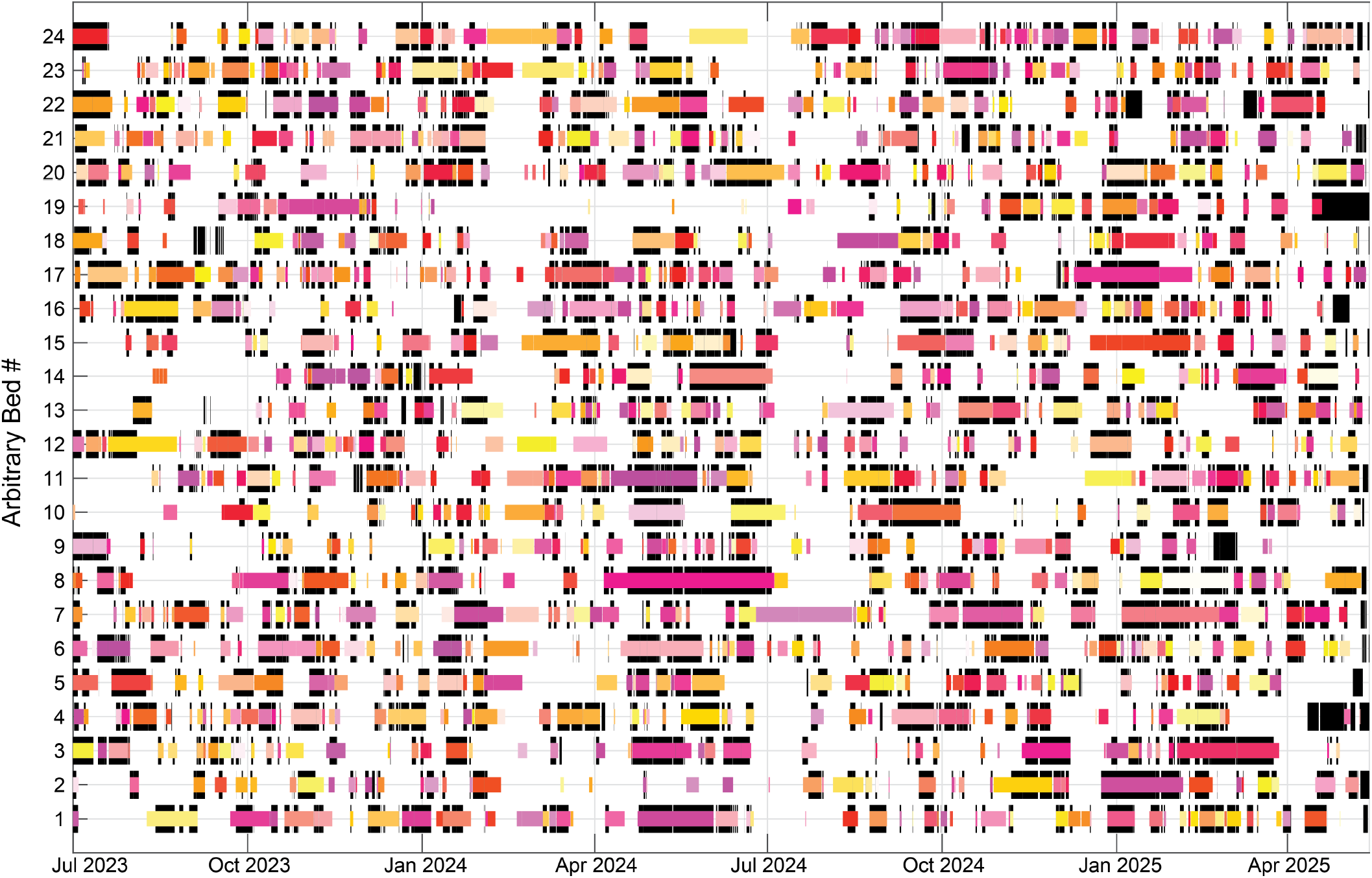
Ventilator Utilization vs Bed Occupancy. Overlap between ventilator waveform data (heavy black lines) and patient bed occupancy from the EHR (narrower colored lines). Each patient is shown in a unique color. EHR data is only received for discharged patients, which explains the ventilator waveforms without corresponding patients from April 2025 onward.

Hamilton’s dual-mode ventilation (adaptive pressure ventilation with controlled mandatory ventilation (APVcmv)) was the most commonly utilized (59.4% of breaths), followed by pressure control (16.6% of breaths) and pressure support (14.2% of breaths). The SCM was fit to each breath. The SCM coefficient of determination (COD) is a measure of fit quality for which we assigned an empirical threshold COD = 0.985 based on visual inspection of the data to identify breaths with signs of respiratory effort. Using this threshold, respiratory effort (or other factors) decreased model fit in 97 million breaths. In the 49 million breaths with accurate fits, the compliance was 35.7 [25.2, 45.3] mL/cmH_2_O and the resistance was 11.4 [9.7, 13.8] cmH_2_O/L/s. Table 2 shows basic descriptors calculated for each breath. These SCM calculated compliance values correlated with clinical values recorded by the RT (Pearson Correlation: 0.86) and could be calculated more frequently; a median of every 0.046 minutes [0.038, 0.056] compared to RT documentation (a median of every 255 minutes [195, 390]). Finally, mechanical power was calculated for every breath from pressure and flow waveforms, with each breath dissipating a median of 8.8 [6.2, 12.5] J/s.

**Table 2:**
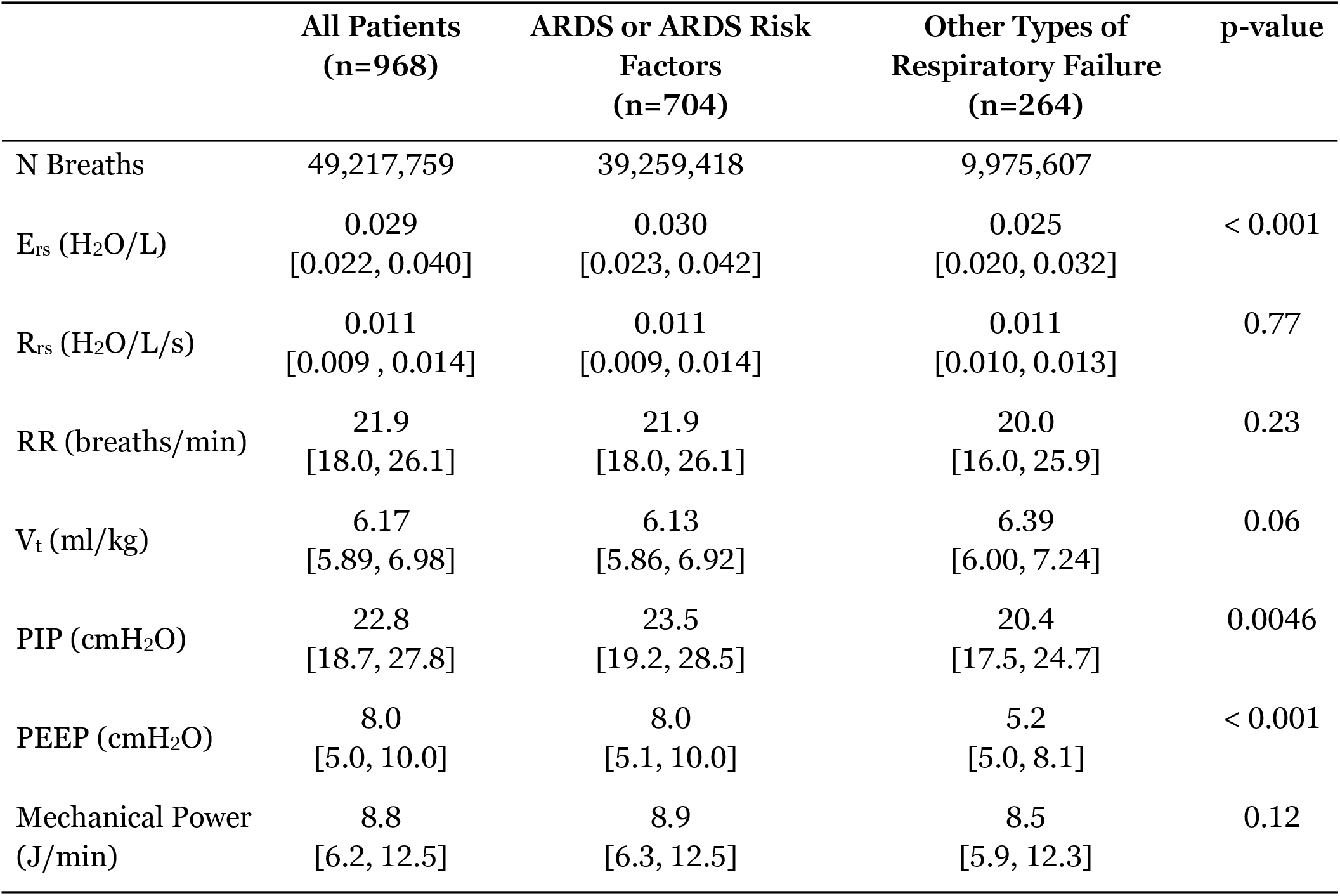
Descriptive Properties of Breaths of in Patients with ARDS or ARDS Risk factors compared to other etiologies of respiratory failure. **E**_**rs**_: respiratory system elastance which is the inverse of compliance, **R**_**rs**_: respiratory system resistance. **RR**: respiratory rate, **V**_**t**_: observed tidal volume, normalized to predicted body weight, **PIP**: peak inspiratory pressure, **PEEP**: positive end expiratory pressure. All reported values are median [25%, 75% inter-quartile range].

|  | **All Patients**<br>**(n=968)** | **ARDS or ARDS Risk**<br>**Factors**<br>**(n=704)** | **Other Types of**<br>**Respiratory Failure**<br>**(n=264)** | **p-value** |
| --- | --- | --- | --- | --- |
| N Breaths | 49,217,759 | 39,259,418 | 9,975,607 |  |
| $E_{rs}$ (H<sub>2</sub>O/L) | 0.029<br>[0.022, 0.040] | 0.030<br>[0.023, 0.042] | 0.025<br>[0.020, 0.032] | < 0.001 |
| $R_{rs}$ (H<sub>2</sub>O/L/s) | 0.011<br>[0.009, 0.014] | 0.011<br>[0.009, 0.014] | 0.011<br>[0.010, 0.013] | 0.77 |
| RR (breaths/min) | 21.9<br>[18.0, 26.1] | 21.9<br>[18.0, 26.1] | 20.0<br>[16.0, 25.9] | 0.23 |
| $V_t$ (ml/kg) | 6.17<br>[5.89, 6.98] | 6.13<br>[5.86, 6.92] | 6.39<br>[6.00, 7.24] | 0.06 |
| PIP (cmH<sub>2</sub>O) | 22.8<br>[18.7, 27.8] | 23.5<br>[19.2, 28.5] | 20.4<br>[17.5, 24.7] | 0.0046 |
| PEEP (cmH<sub>2</sub>O) | 8.0<br>[5.0, 10.0] | 8.0<br>[5.1, 10.0] | 5.2<br>[5.0, 8.1] | < 0.001 |
| Mechanical Power<br>(J/min) | 8.8<br>[6.2, 12.5] | 8.9<br>[6.3, 12.5] | 8.5<br>[5.9, 12.3] | 0.12 |

EHR data was similarly rich, with a median of 8,511 [3,835, 17,040] records per patient. For instance, a total of 12,351 ABGs were collected during these patients’ care, a median of 6 [2, 16] per patient, with a mean pH of 7.37 ± 0.078, P_a_CO_2_ of 37.1 ± 9.1, and P_a_O_2_ of 97.7 ± 27.3. Moreover, an hourly 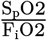 ratio could be easily calculated from documented clinical flow sheet data, yielding 122,000 data points with a mean of 185.7 ± 67.2 and generating a high fidelity measure of respiratory failure severity that is highly correlated with mortality.(41–45) Finally, the Richmond Agitation and Sedation Score (RASS) is documented a median of 75 times per patient [IQR 30-134] and a median score of −0.87 [−1.71, −0.41]. A total of 96% of patients received continuous analgesics with fentanyl drips, and 93% received sedation with propofol drips. A total of 7.2% of patients received neuromuscular blockade (NMB) during their course of mechanical ventilation.

## Discussion

This paper describes a comprehensive data pipeline for integrating continuous ventilator waveform data with detailed EHR data necessary to describe the complex interactions between lung injury, patient effort, ventilator dyssynchrony, sedation, and ventilator mechanics.

The data pipeline and resulting dataset have several unique aspects. First, the dataset captures the patient’s entire hospital course. This facilitates detailed analysis of patient trajectories. Second, we capture both ventilator and EHR data. With this dense data collection, we can define high-fidelity subphenotypes and treatable traits, delineate sophisticated lung state markers, and track intermediate, short-term markers of VILI. This allows us to investigate how clinical interventions affect short-term changes, such as those observed 6 or 12 hours later, as well as classic long-term outcomes, including ventilator length of stay and mortality. Third, the dataset includes numerous time-varying covariates, such as markers of lung physiology, evolving sedation strategies, and temporal changes in the severity of illness. This enables complex time-series analysis that surpasses traditional multivariable adjustments based on variables collected at admission and analyzed in relation to discharge outcomes. Finally, these data include all mechanically ventilated patients in the MICU, a rich and heterogeneous population with causes of acute respiratory failure, including, but not limited to, ARDS.

Moreover, this data pipeline represents a significant multidisciplinary investment. A skilled team of data scientists, biomedical engineers, health system information technology experts, and clinicians was necessary to build, test, and validate the end data. Indeed, without HDC, obtaining EHR data would have been infinitely more difficult. Even with the HDC team in place, building and refining the waveform data pipeline took over 2 years and was only possible with the UCHealth team’s gracious and skilled assistance.

This study has several limitations. First, we record waveform data only in the MICU. While EHR data is available, periods of mechanical ventilation in other units (i.e., emergency department, operating room, time in other ICUs, or during transport) are missing from the waveform data. Expansion to include other departments is feasible in future iterations of our data pipeline and requires simply expanding the Capsule licensing to these units. Transportation periods are generally short — less than 10 minutes — and unlikely to significantly alter a patient’s clinical course. Second, that data is observational, limiting causal analysis. Although the data set is rich, unmeasured confounders are present. Particularly, it is difficult to capture the reasoning behind a given clinical intervention. Third, this dataset is from a single ICU (MICU) at a single hospital, using a single brand of ventilator. Thus, any observations may not be generalizable beyond our institute. Finally, this data is derived from the EHR. Finally, these data incorporate elements of the EHR, such as vital signs and ICD-10 codes, that are manually entered and susceptible to error. Similarly, because data are entered irregularly during clinical care, it is impossible to quantify missingness from the dataset.

Many of these limitations can be overcome by generating large-scale, multi-site ventilator waveforms and EHR datasets containing a spectrum of ventilator types and care practices. This requires first establishing standards for data formatting, structure, and file types. For the EHR, we must determine a set of necessary covariates with defined data types and units. For categorical variables, such as ventilator mode, standard definitions must be established and consistently applied. Definitions related to waveforms will be more challenging. They should start with a common language for breath types (e.g., ventilator dyssynchrony types) that clearly delineates the start and end of every kind of breath. We must then establish a standard methodology for segmenting breaths to enable interoperability between data collected from and analyses performed at multiple sites.

## Conclusion

For the first time, we created a fully automated data pipeline to continuously collect mechanical ventilation waveform data and integrate it with detailed EHR data to generate a unique, high-fidelity dataset. This dataset will facilitate future observational studies to delineate the complex relationships between lung injury, patient effort, sedation, ventilator dyssynchrony, and ventilator mechanics. Moreover, creating such a data pipeline is the first step in leveraging these data streams for real-time clinical utilization, which involves building, testing, and validating predictive models to inform clinical decision-making.

## Data Availability

All data produced in the present study are available upon reasonable request to the authors

## Declarations

### Ethics Approval

This data pipeline and study were approved by the University of Colorado Combined Institutional Review Board (COMIRB, #20-2160).

### Availability of data and materials

The datasets generated and/or analyzed during the current study are not publicly available due to PHI. Nevertheless, a subset of de-identified and time-shifted data is available from the corresponding author upon reasonable request.

### Authors’ contributions

BS contributed to pipeline design and management, data analysis, and manuscript preparation and revision. PS contributed to pipeline design, data analysis, and manuscript preparation and revision. LL contributed to the design and development of the pipeline and to manuscript revision. DA and JS contributed to data analysis, manuscript preparation, and revision.

## Acknowledgements

We would like to acknowledge Corey Montano, Michele Edelman, Michael Kahn, and Bujia Zhang at HDC for their work and assistance in building the HDC component of this data pipeline. Daniel Hoberecht and Tom Libric at UCHealth provided essential contributions to the development, testing, and deployment of the waveform infrastructure.

## Glossary

ADT: Admit, Discharge, Transfer s5
APRV: airway pressure release ventilation 6
APVcmv: adaptive pressure ventilation with controlled mandatory ventilation 7
ARDS: acute respiratory distress syndrome 4, 5, 7, 8
CCTSI: Colorado Clinical and Translational Sciences Institute 7, s2
COD: coefficient of determination 7
COPD: chronic obstructive pulmonary disease 5
EHR: electronic health record 2, 3, 4, 5, 6, 7, 8, 9, 12, s1, s2, s5
HDC: Health Data Compass 6, 8, 10, s5
HIPAA: Health Insurance Portability and Accountability Act 5, 6
HL7: Health Level Seven International 5, 6, s2
I:E ratio: inspiratory to expiratory time ratio 6
ICD-10: International Classification of Diseases, 10th revision 7
ICU: intensive care unit 4, 8, 9
ILD: interstitial lung disease 5
MICU: Medical Intensive Care Unit 2, 5, 6, 8, 9, s5
PEEP: positive end-expiratory pressure 6, 7
PHI: protected health information s5
RASS: Richmond Agitation and Sedation Score 6, 15
SCM: single compartment model 7, 8
TCP: Transmission Control Protocol s2
VILI: ventilator-induced lung injury 4, 8
VM: virtual machine 6, s2

