## Supplemental Appendix for "Developing A Data Pipeline to Quantify Ventilator Waveforms"

### Supplemental Digital Content

Appendix I ........ s2
- Waveform Data Processing ........ s2
- Transforming Waveform Data Streams ........ s2
- Segmenting Continuous Waveforms ........ s2
- Data Interruption Alerts and Pipeline Downtime ........ s3

Appendix II ........ s5
- EHR Integration ........ s5

*APPENDIX I*

*Waveform Data Processing:* Ventilator data are transferred from the ventilator to the Capsule Neuron 3 at a sampling rate of 50 Hz. The ventilator waveform and scalar data (e.g., settings) utilize a separate output connector than the EHR production data (which is sampled at low frequency), with a Transmission Control Protocol (TCP) connection to an on-premise UCHealth-managed VM over the hospital network (VM2 in Figure 1). A PowerShell script captures the incoming TCP packets and appends the HL7 messages to text files named by timestamp, each containing 1,000 HL7 messages. When complete, each timestamped file is uploaded to a secure Azure cloud storage environment maintained by UCHealth. The Azure instance serves as a container for the incoming HL7 text files. These files contain only room numbers (embedded in the Neuron Capsule identification stamp) and timestamps to associate the data with a unique patient. Finally, the raw waveform data (median 2.85 GB/day [IQR 2.3, 3.5]) are automatically downloaded daily to a dedicated research server using a MATLAB function (Matlab 2024, MathWorks, Natick, MA, USA).

All data processing is then performed on a research computational server (Dell R940xa 4U server with 4x Intel Xenon 8268 processors (24 core/48 thread, 2.9 GHz) for a total of 96C/192T; 1.6 TB RAM; 3.5 TB NVMe storage; and a 19 TB SSD array). The CCTSI maintains this system and ensures compliance with healthcare data privacy regulations.

*Transforming Waveform Data Streams:* Waveform data are ingested onto the research server using a MATLAB function that is automatically executed once daily. That function collates a list of waveform files created in the last 21 days (to account for potential missed downloads) on the UCHealth Azure instance, compares it with the local file cache, and downloads any new files to a date-named local directory. For each file, the waveform variables are extracted from the HL7 messages using regular expressions and stored in temporary tables. Each row in these tables includes arrays of 1 to 15 variable values, the bed number, and the HL7 timestamp. Scalar values (such as the set tidal volume) are extracted into a new set of tables that include the variable codes, values, room identifiers, and timestamps. Next, we extract the waveform data, identified by room number, into a new table. Because the precision of the timestamps in the HL7 message is insufficient to accurately reassemble the waveform (which has a 50Hz sampling rate), the HL7 messages are first assembled in order of receipt. We then check for gaps between messages (greater than 0.3 seconds), which indicate data dropout. Periods between gaps define contiguous segments of waveforms (which include many HL7 messages). Within these contiguous segments, times are reassigned to each data point based on the 50 Hz sampling rate. If the assigned time differs from the HL7 timestamp by more than 5 seconds, indicating timing drift, then a new contiguous time block is defined and reprocessed. The per-room output is saved as MATLAB binary files, named according to the download date and time, in a directory tree organized by room number.

*Segmenting Continuous Waveforms:* Segmenting ventilator waveform streams into discrete breaths is relatively simple at fixed respiratory rates in the absence of patient respiratory efforts, but becomes increasingly complicated in clinical settings when patients are making spontaneous respiratory efforts. To automate this process, we developed a rule-based process to identify the start of each breath. This process was validated using a previously collected dataset of 1.7 million breaths, with breath segmentation defined using Hamilton's DataLogger software, which identifies when the ventilator's inspiratory and expiratory valves open and close.(16, 24) The threshold times, volumes, and flow thresholds in our rule-based breath segmentation were empirically determined by manual adjustment to best match that dataset. Visual examples for each rule are depicted in Figure s1.

1. Exclude periods where the ventilator is on and sending data but there are no breaths (e.g., stand-by mode), by excluding periods where the range of pressures is <2 $cmH_2O$ in a 12-second moving window (Figure s1a).
2. Detect the onset of inspiratory flow (the start of a breath) by identifying when the flow rate crosses from less than 5 L/min to greater than 5 L/min. A flow rate threshold of 5 L/min was chosen to minimize false detection of breath initiation due to noise in the flow waveform (Figure s1b)
3. To avoid false starts due to respiratory efforts, we look back from the start of one breath (Breath B) to the beginning of the previous breath (Breath A) to find the time of the last instance of expiratory flow (flow <−10 L/min, denoted as $t_{neg}$) If there is no expiratory flow since the last instance of inspiratory flow in Breath A (flow >20 L/min, denoted as $t_{pos}$), or the start of Breath A if there is no flow > 20 L/min, then Breath B is merged with Breath A. These periods are indicative of dyssynchrony and merged with the preceding breath (Figure s1c).
4. At each breath initiation, the ventilator resets the reported volume to null. When true end-expiratory volume is not zero and/or flow conditions are not met, steps 2 and 3 may miss breath initiation (mainly from dyssynchrony). To avoid missing these breaths, we take the time derivative of ventilator-reported volume and find time points where that calculated flow and the reported flow differ by >100 L/min and the volume is <50 mL. These points are indicative of the ventilator resetting the volume to zero at the beginning of a delivered breath (Figure s1d). Additionally, breaths with a ventilator-reported volume greater than 100 mL at the start of the breath are merged with the prior breath because the ventilator resets the volume at the beginning of each breath.
5. Every 10 minutes, the Hamilton ventilators re-zero the internal pressure transducer that is responsible for flow measurement. This is a necessary action to prevent volume drift in the data. However, during this approximately 0.5-second period, the reported pressure, flow, and volume are all zero. Any breaths that intersect with these periods are excluded from analysis (Figure s1e).
6. Breaths shorter than 0.4 seconds (corresponding to a respiratory rate > 150 breaths/min) are merged with the prior breath because that rate is not physiologically compatible with adult subjects. This is generally seen with suctioning. Breaths longer than 15 s are also excluded from the analysis.

The resulting breath segmentations (breath numbers) are stored alongside pressure, flow, and volume in a time series table. The segmented breaths are also stored in a data structure indexed by breath number since filtering the table by breath index can be slow for long recordings.

*Data Interruption Alerts and Pipeline Downtime:* A separate PowerShell script running on the research server checks message frequency twice daily and notifies study personnel if the thresholds are not met. The script looks back 14 hours in the Azure storage and checks that an average of >30 files per hour (>30000 HL7 messages) have been received and that the maximum time between messages is <10 minutes. In addition, a summary notification detailing message volume (number of messages and GB of data), the standard deviation of message frequency, and other details is automatically sent twice daily to provide a more nuanced view of data volume and pipeline integrity. These automated notifications have been key to quickly identifying outages and maintaining the pipeline infrastructure.

Figure 2 shows pipeline outages (red) that myriad reasons, including software updates, connectivity issues between the VMs, and system restarts, have caused. The improvements in uptime from October 2024 onward are primarily due to the development of a robust system that detects and automatically restores network disconnections between the Capsule server (Virtual Machine 1) and Virtual Machine 2 that bundles the HL7s into text files and uploads them to Azure (Figure 1). This, for example, avoids manual intervention and data loss when the Capsule VM is restarted for scheduled updates.

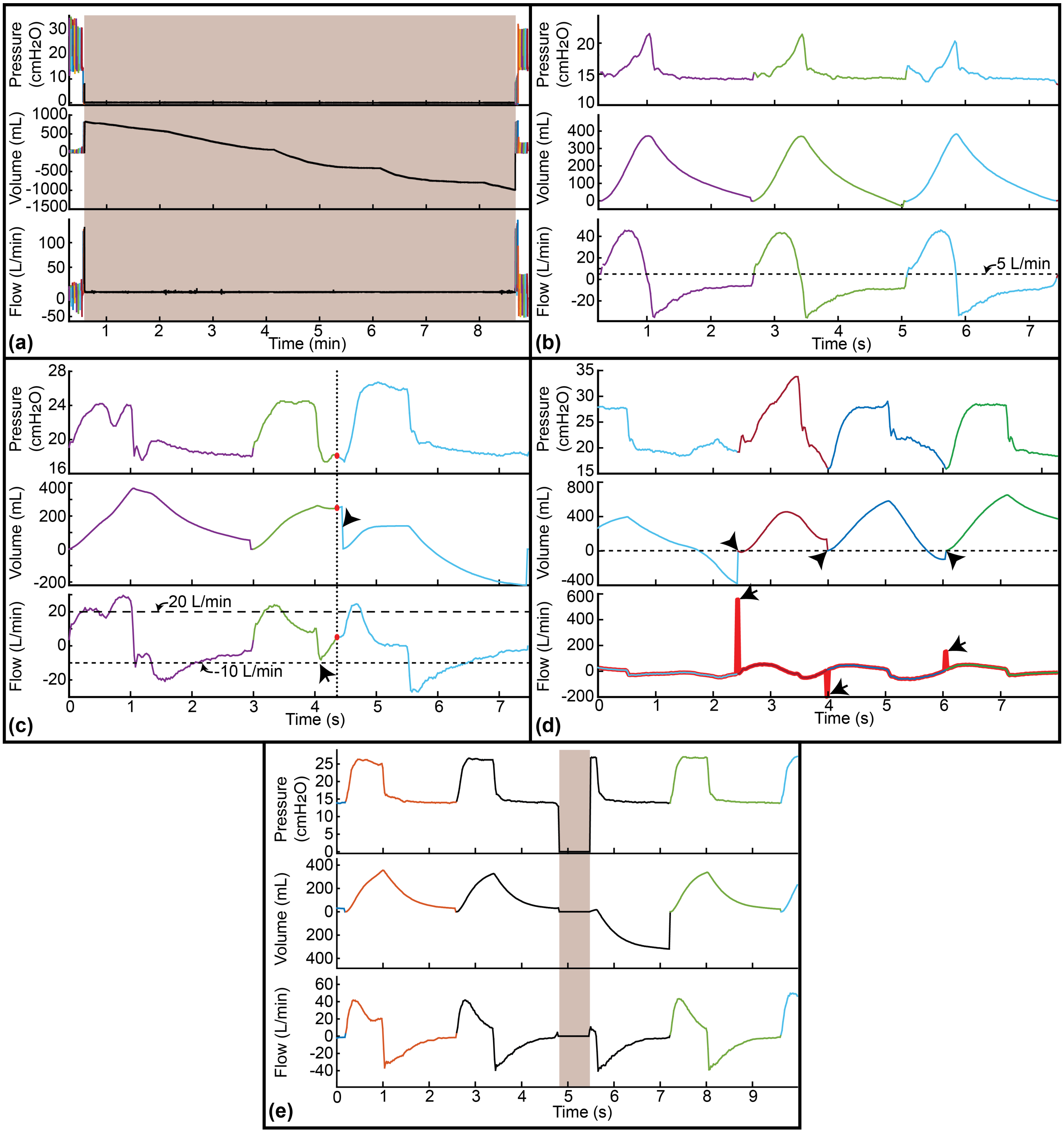


**Figure s1: Waveform Segregation:** A rule-based approach was taken to overcome the active breathing by the patient. In all panels, black lines indicate data that is excluded from further analysis, and colors indicate segmented breaths. **a) Ventilator inactive.** Periods where the ventilator is sending data but there are no breaths (orange shaded area) are eliminated based on the pressure swings being < 2 cmH2O in a 12-second moving window. **b) Breath detection by flow crossing.** Time points where the flow crosses from below to above 5 L/min (dashed line) are demarcated as breath start points. **c) Insufficient negative flow in prior breath.** Breaths start (red circle) where expiratory flow <−10 L/min (short dashed line) does not exist in the prior breath (green) or does not exist after an inspiratory flow > 20 L/min (long dashed line) are merged with the prior breath. Here, the light blue and green lines would be merged into a single breath. Note that the arrowhead indicates a volume reset by the ventilator that would be later identified as a breath start (e.g., panel d). **d) Ventilator volume reset at breath start.** At the initiation of a new breath, the Hamilton ventilator re-zeros the reported volume (arrows). These breath-start events are identified as time points (arrows) where the time derivative of the volume (a flow, heavy red line) differs from the reported flow by > 100 L/min and the volume is < 50 mL (due to re-zeroing, arrowheads). **e) Ventilator recalibration.** The Hamilton ventilator re-zeros the flow sensor at 10-minute intervals (orange shading), and during this time, pressure, flow, and volume are reported as zero. Breaths that intersect this ½-second period are excluded from analysis.

*Appendix II*

*EHR Integration:* The collected ventilator waveform data rely on a combination of unique dates, times, and bed numbers to identify distinct patient encounters. Using custom structured query language (SQL) scripts, HDC identifies patients who were mechanically ventilated in the MICU during the study timeframe. This identifies unique patient hospitalizations and assigns arbitrary, de-identified patient encounter identifiers to track patient data. Consequently, the EHR data request provides specified variables for all patients who receive mechanical ventilation in the MICU during a given hospital stay, linked by these arbitrary identifiers. For each patient, this yields a set of data tables that include ventilator waveforms (from our pipeline) and HDC-provided laboratory test results, physiological measurements, medications, demographic details, in-hospital transfers, outcomes, and other relevant information. EHR data delivered by HDC are de-identified, except for date and time stamps and room numbers, which are needed for waveform linkage. To limit the transfer of protected health information (PHI), the names, medical record numbers, visit numbers, social security numbers, addresses, free-text values, and other sensitive PHI are not included in the data delivered to our research server.

EHR data are delivered to our research server on request. The research team links the ventilator waveform data to the EHR using patients' bed occupancy in the HDC-provided Admit, Discharge, Transfer (ADT) tables, which chronicle the patient's physical location over time. That linkage is established for each patient by identifying temporal overlap between patient bed occupancy and the ventilator waveforms linked to that bed. This script generates a simple diagram to visualize the overlap, ensuring accurate data linkage and identifying periods of missing data (see Figure 3). During software development, we conducted an extensive manual review of the patient-ventilator interface using ADT tables and EHR ventilator data. We also conducted an algorithmic comparison of, for example, EHR-reported and waveform-observed PEEP to assure room and timestamp alignment.
